# Explanatory factors for Ethnic inequalities in Multimorbidity; findings from pooled Health Survey for England 2011-2018

**DOI:** 10.1101/2022.10.03.22280637

**Authors:** Linda Ng Fat, Jennifer S Mindell, Logan Manikam, Shaun Scholes

## Abstract

**Background:** Social-economic factors and health behaviours may be driving variation in ethnic health inequalities in multimorbidity including among distinct ethnic groups.

**Methods:** Using the cross-sectional nationally-representative Health Surveys for England 2011-2018 (N=54,438, aged 16+), we carried out multivariable logistic regression on the odds of having general multimorbidity (≥2 longstanding conditions) by ethnicity (British White (reference group), White Irish, Other White, Indian, Pakistani, Bangladeshi, Chinese, African, Caribbean, White Mixed, Other Mixed, Other), adjusting for age, sex, education, area-deprivation, smoking status, and survey year. This was repeated for cardiovascular multimorbidity (N=37,148, aged 40+: having ≥2 of the following: doctor-diagnosed diabetes or hypertension, heart attack or stroke) and multiple cardiometabolic risk biomarkers (HbA1c ≥6.5%, raised blood pressure, total cholesterol ≥5mmol/L).

**Results:** 20% of adults had general multimorbidity. In fully-adjusted models, compared with the White British majority, Other White (Odds Ratio (OR)=0.63 (95% confidence interval=(0.53-0.74)), Chinese (OR=0.58 (0.36-0.93)), and African adults (OR=0.54 (0.42-0.69)), had lower odds of general multimorbidity. Among adults aged 40+, Pakistani (OR=1.27 (0.97-1.66) p=0.080) and Bangladeshi (OR=1.75 (1.16-2.65)) had increased odds, and African adults had decreased odds (OR=0.63 (0.47-0.83)) of general multimorbidity. Risk of cardiovascular multimorbidity was higher among Indian (OR=3.31 (2.56-4.28)), Pakistani (OR=3.48 (2.52-4.80)), Bangladeshi (OR=3.67 (1.98-6.78)), African (OR=1.61 (1.05-2.47)), Caribbean (OR=2.18 (1.59-2.99)) and White Mixed (OR=1.98 (1.14-3.44)) adults. Indian adults were also at risk of having multiple cardiometabolic risk biomarkers.

**Conclusion:** Ethnic inequalities in multimorbidity are independent of socioeconomic factors. Ethnic minority groups are particularly at risk of cardiovascular multimorbidity, which may be exacerbated by poorer management of cardiometabolic risk factors.

**What is already known on this topic:**

- Studies have found ethnic inequalities in multimorbidity among broad ethnic groups; however, it is not known whether lower socioeconomic status or health behaviours are driving this inequality in England, nor whether there are differences between distinct ethnic groups.

**What this study adds:**

- Ethnic inequalities in multimorbidity in a nationally-representative sample in England vary between older and younger adults, distinct ethnic groups, and are independent of socioeconomic status, smoking and obesity.
- African, Caribbean, Bangladeshi, Indian, Pakistani and White mixed adults aged 40 and over had higher risk of self-reported cardiovascular multimorbidity compared with British White adults. Indian adults were also at risk of multiple uncontrolled cardiometabolic risk biomarkers.
- African, Chinese, Other White and Indian adults (<40 years) had lower risk of general multimorbidity compared with the British White majority, suggesting a healthy migrant effect or differences in interpretation of chronic conditions.

**How this study might affect research, practice or policy:**

- Further research is needed to explore whether underdiagnosis and/or poorer management of cardiometabolic risk factors among ethnic groups may be driving factors behind inequalities in self-reported cardiovascular multimorbidity.

## Background

Multimorbidity is increasing, given an ageing population, affecting individuals’ quality of life and increasing mortality,(1,2). Understanding the factors related to different levels of multimorbidity in ethnic groups could be used to prevent harm and reduce ethnic inequalities. Non-white ethnic groups were disproportionately affected with Covid-19, with multimorbidity potentially being one contributing factor. It was found that compared with the white population, non-white ethnicities with multimorbidity had almost triple the risk of having Covid-19(3).

Ethnic health inequalities in multimorbidity, including cardiovascular multimorbidity, exist(4– 9). A study in East London found the Black and South Asian population had higher rates of cardiovascular multimorbidity than the white majority(4). Studies of ethnic health inequalities in multimorbidity in England often use GP records, which may not reflect differences in a national population sample,(4,5). Furthermore, studies in England have not comprehensively explored what may be accounting for some of the health differences, such as socioeconomic differences. Deprivation and lower socioeconomic status could increase the risk of multimorbidity,(6–8,10–12). Behavioural risk-factors could also confound the relationship. Smoking and obesity were found to be risk-factors for multimorbidity,(7,10,13), and may have a stronger effect than deprivation and ethnicity,(7,13). Smoking and obesity were found to vary in The Health Survey for England 2011-2019,(14).

Furthermore, broad ethnic groups such as ‘South Asian’ and ‘Black’ may mask heterogeneity within groups, for example among Indian versus Pakistani/Bangladeshi, and Black African versus Black Caribbean groups,(15). The aims of this study were to examine how risk of multimorbidity including cardiovascular multimorbidity varied among distinct ethnic groups, and whether this relationship was explained by difference in socioeconomic status or behavioural risk factors for cardiovascular diseases, namely smoking and obesity.

## Methods

### Data Sources

The Health Survey for England (HSE) is a cross-sectional, nationally-representative survey of the population occurring annually. Data is collected via new multistage stratified probability sampling each year, as described elsewhere(16). This study uses data spanning from 2011 to 2018, to maximise the number of participants from different ethnic groups. Self-reported data were collected from an initial face-to-face interview conducted by trained interviewers using computer-assisted personal interviewing (CAPI). The interviewers also measured height and weight. At the second stage, further biophysical measurements were collected at a nurse visit using standard protocols,(17). All data collection took place within participants’ homes. Response rates have gradually declined over the period, e.g., from 59% in 2011 to 54% in 2018 for the face-to-face interview.

### General and cardiovascular multimorbidity

This study examines ethnic group differences in two types of multimorbidity: general multimorbidity and cardiovascular multimorbidity, using information provided by participants at the interview stage. General multimorbidity was derived through self-reported responses to a question on longstanding conditions: whether the participant had any longstanding physical or mental health conditions or illnesses expected to last over a period of time (2011) or 12 months or more (2012-2018). Participants were then asked to list up to six conditions that affected them. Responses were recoded to indicate the number of longstanding conditions (none/one/two or more). Participants having two or more longstanding conditions were classified as having general multimorbidity.

Self-reported doctor-diagnosed cardiovascular multimorbidity was defined as having two or more of the following conditions: reporting having doctor-diagnosed diabetes or high blood pressure (in response to specific questions on whether participants were ever told by a doctor they had diabetes/high blood pressure) or reporting stroke or heart disease when responding to the aforementioned follow-up question to the longstanding illness question. We included doctor-diagnosed hypertension following the example of Mathur et al.(4), although other definitions do not include hypertension,(18).

### Multiple cardiometabolic risk biomarkers

Using objective health examination measures, we also examined differences by ethnicity in multiple cardiometabolic risk biomarkers. Cardiometabolic biomarkers were treated separately from doctor-diagnosed conditions (cardiovascular multimorbidity) to identify uncontrolled risk factors. Having multiple cardiometabolic risk biomarkers was defined as two or more of the following: raised glycated haemoglobin (HbA_1c_ levels ≥6.5%); raised blood pressure (systolic blood pressure ≥140mmHg and/or diastolic blood pressure ≥90mmHg); and/or raised total cholesterol level (≥5mmol/L), suggesting uncontrolled risk-factors or disease such as diabetes or hypertension. Further details on data collection during the nurse visit have been published elsewhere,(15).

### Ethnicity and other covariates

Ethnicity was self-reported at the main interview and grouped in the following way: White British, White Irish, Other White, Indian, Pakistani, Bangladeshi, Chinese, African, Caribbean, White Mixed, Other Mixed, Other.

Other covariates included the Index of Multiple Deprivation, a standard measure of area deprivation covering different social and economic dimensions, assigned based on participants’ residence,(17), grouped into quintiles. Highest educational qualification was self-reported and grouped into degree or higher; lower than degree; no qualifications. Self-reported smoking status was grouped into never; former; current smoker. BMI categories were based on height and weight measurements of the participants performed by the trained interviewer at the main interview. Detailed information on collection can be found elsewhere,(17).

### Statistical analyses

Complete case analyses were used and the sample was limited to (i) adults aged 16+ for general multimorbidity (N=54,438), (ii) adults aged 40+ for self-reported cardiovascular multimorbidity (N= 37,148), and (iii) adults aged 16+ with biological measurements for multiple cardiometabolic biological risk factors (N=24,203). The complete case approach resulted in a reduction in sample size of around 17.5%; this was predominately driven by missing data for BMI, which required physical measurements of height and weight (17.0%). Few participants (<0.5%) had missing responses on key variables: general multimorbidity (n=46), cardiovascular multimorbidity (n=7), and ethnicity (n=220). Missing data on biomarkers was high due to not all the sample providing blood samples (59.8%). We accounted for non-response weighting, specific to non-response at the interview or blood samples for biological risk factors were applied to account for this, and the complex survey design (geographical clustering of participants) in al analyses.

Characteristics of ethnic groups were explored and chi-squared used to test for bivariate associations. Logistic regression was carried out on the risk of having multimorbidity for each of the three definitions of multimorbidity. Since longstanding conditions are broader, and multimorbidity is not restricted to older participants,(8,19), analyses of general multimorbidity focused on all adults. However, we also stratify results by aged under 40 and 40 and over. A cut-off of age 40 has been used in other large scale studies,(20), and sample sizes among some of the ethnic groups were too small if limiting to an older sample (e.g. <100, if based on adults aged 65 and over).

To explore whether the relationship between ethnicity and multimorbidity may be confounded by socioeconomic status or behavioural risk factors, variables were added in the following way, with changes in odds ratios examined to explore the effect of adjusting for each additional potential confounder.

Model 1: Sex + Age in years

Model 2: Model 1 + Area Deprivation

Model 3: Model 1 + Educational qualifications

Model 4: Model 1 + Smoking status

Model 5: Model 1 + BMI category

Model 6: All covariates + survey year

As a sensitivity analyses, we replicated the final models on a sample including participants without BMI measurements (without adjusting for BMI), resulting in fewer missing cases (1%). Analyses were conducted in Stata 17,(21).

## Results

Chinese, Indian and Other White adults had the highest proportion of adults with the highest educational qualifications (53%-44%), whereas Bangladeshi, White Irish, Pakistani, and Caribbean had the highest proportions with no qualifications (29%-25%) (Table 1). Over two-fifths of adults in the Bangladeshi, Pakistani, African and Caribbean groups lived in the most deprived areas; this proportion was lowest for White British adults (15%).

**Table 1.** Characteristics of ethnic groups (column %), Health Survey for England 2011-2018.

|  | White<br>British | White<br>Irish | Other<br>White | Indian | Pakistani | Bangladeshi | Chinese | African | Caribbean | White<br>Mixed | Other<br>Mixed | Other | Total | p-value |
| --- | --- | --- | --- | --- | --- | --- | --- | --- | --- | --- | --- | --- | --- | --- |
| **Sample size (N)** | 44,922 | 440 | 2,876 | 1,382 | 859 | 368 | 278 | 804 | 496 | 477 | 268 | 1,268 | 54,438 |  |
| **Mean age (years)** | 48.8 | 53.4 | 37.6 | 40.6 | 36.8 | 36.5 | 36.2 | 37.3 | 48.6 | 34.1 | 38.8 | 39.8 | 46.8 |  |
| *standard deviation* | 19 | 17.2 | 12.5 | 14.1 | 13 | 12.4 | 13.9 | 13.1 | 17.4 | 13.4 | 14 | 14.6 | 18.5 |  |
| **Female** | 50 | 47 | 52 | 48 | 50 | 49 | 53 | 52 | 56 | 49 | 51 | 51 | 50 | 0.103 |
| **Highest Educational qualification** |  |  |  |  |  |  |  |  |  |  |  |  |  |  |
| Degree or higher | 25 | 39 | 44 | 45 | 29 | 26 | 53 | 37 | 23 | 31 | 40 | 41 | 28 |  |
| Lower than degree | 57 | 33 | 32 | 38 | 45 | 45 | 32 | 48 | 52 | 54 | 51 | 43 | 53 |  |
| No qualifications | 18 | 28 | 25 | 17 | 26 | 29 | 14 | 15 | 25 | 15 | 10 | 17 | 19 | <0.001 |
| **Index of Multiple Deprivation** |  |  |  |  |  |  |  |  |  |  |  |  |  |  |
| Least deprived | 23 | 18 | 13 | 15 | 5 | 3 | 19 | 5 | 8 | 12 | 11 | 13 | 20 |  |
| 2<sup>nd</sup> | 23 | 15 | 16 | 17 | 7 | 7 | 15 | 9 | 7 | 16 | 18 | 16 | 21 |  |
| 3<sup>rd</sup> | 22 | 20 | 20 | 20 | 11 | 8 | 24 | 12 | 14 | 17 | 24 | 17 | 21 |  |
| 4<sup>th</sup> | 18 | 26 | 27 | 25 | 29 | 26 | 27 | 31 | 28 | 25 | 27 | 25 | 20 |  |
| Most deprived | 15 | 21 | 24 | 23 | 49 | 56 | 15 | 43 | 43 | 30 | 20 | 29 | 19 | <0.001 |
| **Smoking status** |  |  |  |  |  |  |  |  |  |  |  |  |  |  |
| Current smoker | 19 | 20 | 27 | 8 | 15 | 18 | 10 | 7 | 21 | 29 | 17 | 12 | 19 |  |
| Ex-smoker | 27 | 34 | 22 | 8 | 8 | 8 | 7 | 8 | 18 | 17 | 17 | 12 | 25 |  |
| Never smoker | 54 | 47 | 52 | 84 | 77 | 74 | 83 | 85 | 61 | 54 | 67 | 76 | 57 | <0.001 |
| **Body Mass Index (BMI) category** |  |  |  |  |  |  |  |  |  |  |  |  |  |  |
| Not overweight | 36 | 38 | 48 | 44 | 36 | 44 | 74 | 36 | 29 | 46 | 44 | 43 | 38 |  |
| Overweight | 37 | 35 | 33 | 38 | 37 | 42 | 22 | 33 | 32 | 31 | 35 | 35 | 36 |  |
| Obese | 27 | 27 | 20 | 18 | 27 | 15 | 3 | 31 | 39 | 23 | 21 | 22 | 26 | <0.001 |
| **Number of longstanding conditions** |  |  |  |  |  |  |  |  |  |  |  |  |  |  |
| None | 58 | 57 | 76 | 70 | 71 | 67 | 86 | 77 | 55 | 68 | 72 | 71 | 61 |  |
| One | 21 | 17 | 14 | 17 | 15 | 16 | 7 | 14 | 22 | 17 | 15 | 16 | 20 |  |
| Two or more | 22 | 25 | 10 | 13 | 15 | 17 | 7 | 9 | 24 | 15 | 13 | 13 | 20 | <0.001 |
| **Number of longstanding conditions (aged 40 and over, N=37,135)** |  |  |  |  |  |  |  |  |  |  |  |  |  |  |
| None | 49 | 50 | 63 | 55 | 53 | 49 | 79 | 67 | 46 | 51 | 64 | 56 | 51 |  |
| One | 23 | 20 | 17 | 22 | 19 | 18 | 10 | 17 | 24 | 24 | 16 | 23 | 22 |  |
| Two or more | 28 | 31 | 20 | 23 | 29 | 33 | 12 | 16 | 30 | 25 | 20 | 21 | 27 | <0.001 |
| **Number of cardiovascular conditions (aged 40 and over, N= 37,148)<sup>1</sup>** |  |  |  |  |  |  |  |  |  |  |  |  |  |  |
| None | 62 | 59 | 75 | 62 | 62 | 64 | 81 | 64 | 51 | 69 | 67 | 65 | 63 |  |
| One | 31 | 32 | 21 | 26 | 23 | 22 | 15 | 28 | 33 | 22 | 29 | 26 | 30 |  |
| Two or more | 7 | 9 | 4 | 12 | 15 | 15 | 4 | 7 | 16 | 9 | 4 | 9 | 7 | <0.001 |
| **Cardiometabolic risk biomarkers (aged 16 and over with blood samples (aged 16 and over, N=24,203)<sup>2</sup>** |  |  |  |  |  |  |  |  |  |  |  |  |  |  |
| None | 38 | 31 | 53 | 44 | 44 | 45 | 58 | 53 | 36 | 56 | 54 | 47 | 40 |  |
| One | 47 | 52 | 40 | 42 | 48 | 47 | 35 | 38 | 45 | 39 | 39 | 43 | 46 |  |
| Two or more | 14 | 17 | 7 | 15 | 8 | 9 | 7 | 10 | 19 | 6 | 7 | 10 | 14 | <0.001 |

<sup>1</sup> Doctor-diagnosed hypertension, diabetes or self-reported stroke or heart attack; <sup>2</sup> Raised blood pressure, raised glycated haemoglobin or raised total cholesterol;

Obesity was highest among Caribbean and African adults (39%-31%); and was lowest among Chinese, Bangladeshi and Indian adults (3%-18%). The proportion of never smoking was highest among African, Indian, Chinese, Pakistani and Bangladeshi adults (85%-74%) and was lowest among White Irish, White and White mixed adults (47%-54%).

20% of all adults had general multimorbidity (27% of adults aged 40+); this was higher among White Irish and Caribbean adults (24% and 25%, respectively), and lowest among Chinese and African adults (7% and 9%) (Table 2). 7% of adults aged 40+ had cardiovascular multimorbidity. This was highest among Caribbean, Pakistani and Bangladeshi adults (15%) and lowest among Chinese and Other White adults (4%).

**Table 2.** Characteristics of those with single and multiple limiting longstanding illness conditions (column %), HSE 2011-2018.

|  | None | One | Two or more | Total | p-value |
| --- | --- | --- | --- | --- | --- |
| Sample size (N) | 31,233 | 11,087 | 12,118 | 54,438 |  |
| **Mean age (years)** | 41.9 | 50 | 58.3 | 46.8 |  |
| *Standard deviation* | 16.5 | 18.5 | 18.2 | 18.5 |  |
| **Female** | 49 | 52 | 54 | 50 | <0.001 |
| **Highest educational qualification** |  |  |  |  |  |
| Degree or higher | 31 | 27 | 19 | 28 |  |
| Lower than degree | 55 | 54 | 50 | 53 |  |
| No qualifications | 14 | 20 | 32 | 19 | <0.001 |
| **Index of multiple deprivation** |  |  |  |  |  |
| Least deprived quintile | 21 | 21 | 17 | 20 |  |
| 2<sup>nd</sup> quintile | 21 | 21 | 20 | 21 |  |
| 3<sup>rd</sup> quintile | 21 | 22 | 21 | 21 |  |
| 4<sup>th</sup> quintile | 19 | 19 | 21 | 20 |  |
| Most deprived quintile | 18 | 17 | 22 | 19 | <0.001 |
| **Smoking status** |  |  |  |  |  |
| Current | 18 | 19 | 19 | 19 |  |
| Ex-regular | 21 | 28 | 34 | 25 |  |
| Never regular | 61 | 53 | 47 | 57 | <0.001 |
| **Body Mass Index (BMI) category** |  |  |  |  |  |
| Not overweight | 43 | 34 | 25 | 38 |  |
| Overweight | 37 | 37 | 35 | 36 |  |
| Obese | 21 | 29 | 40 | 26 | <0.001 |
| **Number of cardiovascular conditions (aged 40+, row %)<sup>1</sup>** |  |  |  |  |  |
| None | 63 | 21 | 17 | 100 |  |
| One condition | 36 | 26 | 39 | 100 |  |
| Two or more conditions | 9 | 18 | 73 | 100 | <0.001 |
| **Year (row %)** |  |  |  |  |  |
| 2011 | 61 | 21 | 18 | 100 |  |
| 2012 | 63 | 19 | 18 | 100 |  |
| 2013 | 62 | 19 | 20 | 100 |  |
| 2014 | 61 | 19 | 20 | 100 |  |
| 2015 | 62 | 20 | 19 | 100 |  |
| 2016 | 60 | 20 | 21 | 100 |  |
| 2017 | 58 | 20 | 22 | 100 |  |
| 2018 | 59 | 19 | 22 | 100 |  |
| 2011-2018 | 61 | 20 | 20 | 100 | <0.001 |

<sup>1</sup> Doctor-diagnosed hypertension, diabetes or self-reported stroke or heart attack

Among those aged 40+ with cardiovascular multimorbidity, the combination of diabetes and hypertension was the most common dyad (70%) followed by hypertension and heart attack (12%) (Figure S1). 14% of adults aged 16+ had multiple cardiometabolic risk biomarkers; this varied by ethnicity, being highest among White Irish, Caribbean and Indian adults (15%-19%). This largely comprised the combination of raised blood pressure and raised cholesterol (76%), followed by raised cholesterol and raised glycated haemoglobin (9%)

Those with general multimorbidity were more likely to have no qualifications than those with no longstanding conditions, lived in the most deprived areas, were obese or were a current or former smoker (p<0.001) (Table 2). Among those classified as having cardiovascular multimorbidity (aged 40+), 73% were classified as having general multimorbidity (p<0.001). The prevalence of general multimorbidity increased steadily over the period, from 18% in 2011 to 22% in 2018 (p<0.001).

In age-sex adjusted models, compared with White British adults, Other White (OR=0.66 95% Confidence Interval (0.57-0.77)), Indian (0.78, (0.65-0.93)), Chinese (0.42 (0.26-0.68)), African (0.63 (0.50-0.80)) and Other adults (0.76, (0.63-0.91)) had lower odds of general multimorbidity. The odds for other ethnic groups were not statistically significant although Bangladeshi adults had higher odds (1.34, (0.98, 1.84), p=0.069) (Figure 1, Table S1). In fully adjusted models, the results remained significant only for Other White (0.63 (0.53-0.74)), Chinese (0.58, (0.36-0.93)) and African adults (0.54 (0.42-0.69)).

**Figure 1.**
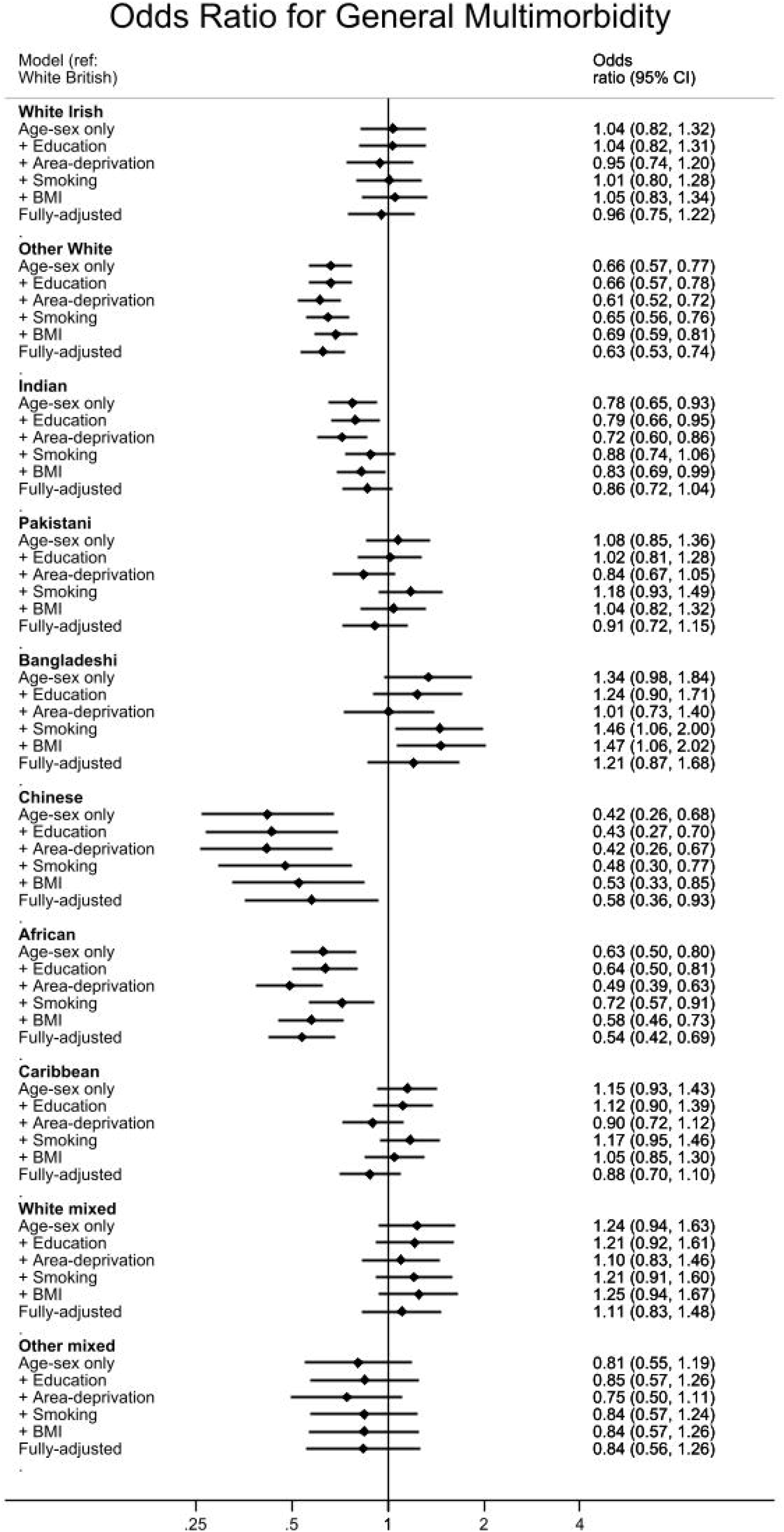
General multimorbidity

In models adjusting for single additional covariates, significant effect sizes of reduced risk were strengthened after adjusting for area deprivation, whilst adjusting for current smoking status reduced effect sizes. Bangladeshi adults had higher odds of general multimorbidity after adjusting for smoking status or BMI status, however, this association was not significant after fully adjusting for all covariates.

Among adults aged under 40, in fully adjusted models, African (0.37 (0.21-0.62)), Indian (0.37 (0.24-0.57)), Other White (0.41 (0.29-0.58)), Pakistani (0.51 (0.32-0.78)) and Other adults (0.50 (0.34-0.75)), had reduced odds of general multimorbidity compared with White British adults (Table S2). Among adults aged 40 and over in fully adjusted models (Table S3), compared with White British adults, Bangladeshi (1.27 (0.97-1.66) p=0.080) and Pakistani adults (1.75 (1.16-2.65)) had higher odds of having general multimorbidity, whereas Other White (0.81, (0.68-0.96)), Chinese adults (0.62 (0.36-1.07, p=0.086)) and African adults had lower odds (0.63, (0.47-0. 83)).

Figure 2 (Table S4) presents odds ratios for cardiovascular multimorbidity among adults aged 40+. In age-sex adjusted models, compared with White British adults, Pakistani (4.01 (2.94-5.46)), Bangladeshi (3.73 (2.15-6.47)), Caribbean (2.93 (2.15-3.98)), Indian (2.52 (1.97-3.24)), White mixed (2.23 (1.32-3.76)), Other (1.93 (1.46-2.54)), and African adults (1.86 (1.23-2.81)) had higher odds of having cardiovascular multimorbidity. In fully adjusted models, the relationship remained significant for all ethnicities. The effect sizes were attenuated for Caribbean (2.18 (1.58-2.99)), African (1.61 (1.05-2.47)), Pakistani (3.48 (2.52-4.80)), White Mixed (1.98 (1.14-3.44)), and Bangladeshi (3.67 (1.98-6.78)) adults and strengthened for Indian (3.31 (2.56-4.28)) and Other adults (2.12 (1.60-2.82)).

**Figure 2.**
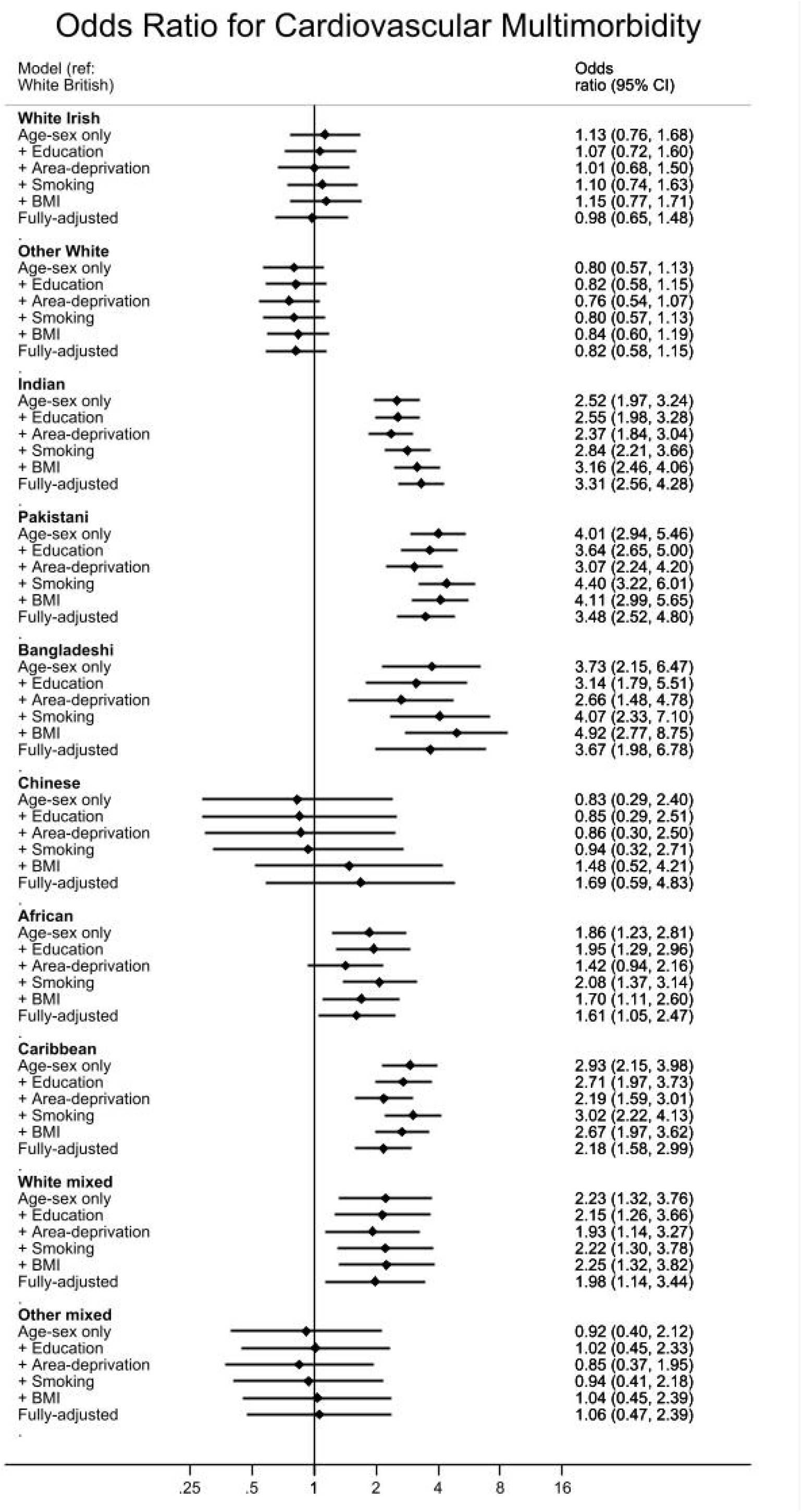
Cardiovascular multimorbidity

Broadly, introducing area-deprivation reduced effect sizes for all significant effects, whereas adjusting for smoking strengthened the effects (with the exception of White Mixed adults). Adjusting for BMI status had a differential effect, slightly attenuating effect sizes for African and Caribbean adults whilst strengthening effect sizes for Bangladeshi, Indian, Other Mixed and Pakistani adults.

In sex-age adjusted models, among adults aged 16+, compared with White British adults, Indian (1.54 (1.17-2.03)) and Caribbean adults (1.46 (0.98-2.19), p=0.064) had greater odds of multiple risk biomarkers (Figure 3). In fully adjusted models, the results were strengthened for Indian adults (1.81 (1.36-2.41)) and were no longer significant for Caribbean adults.

**Figure 3.**
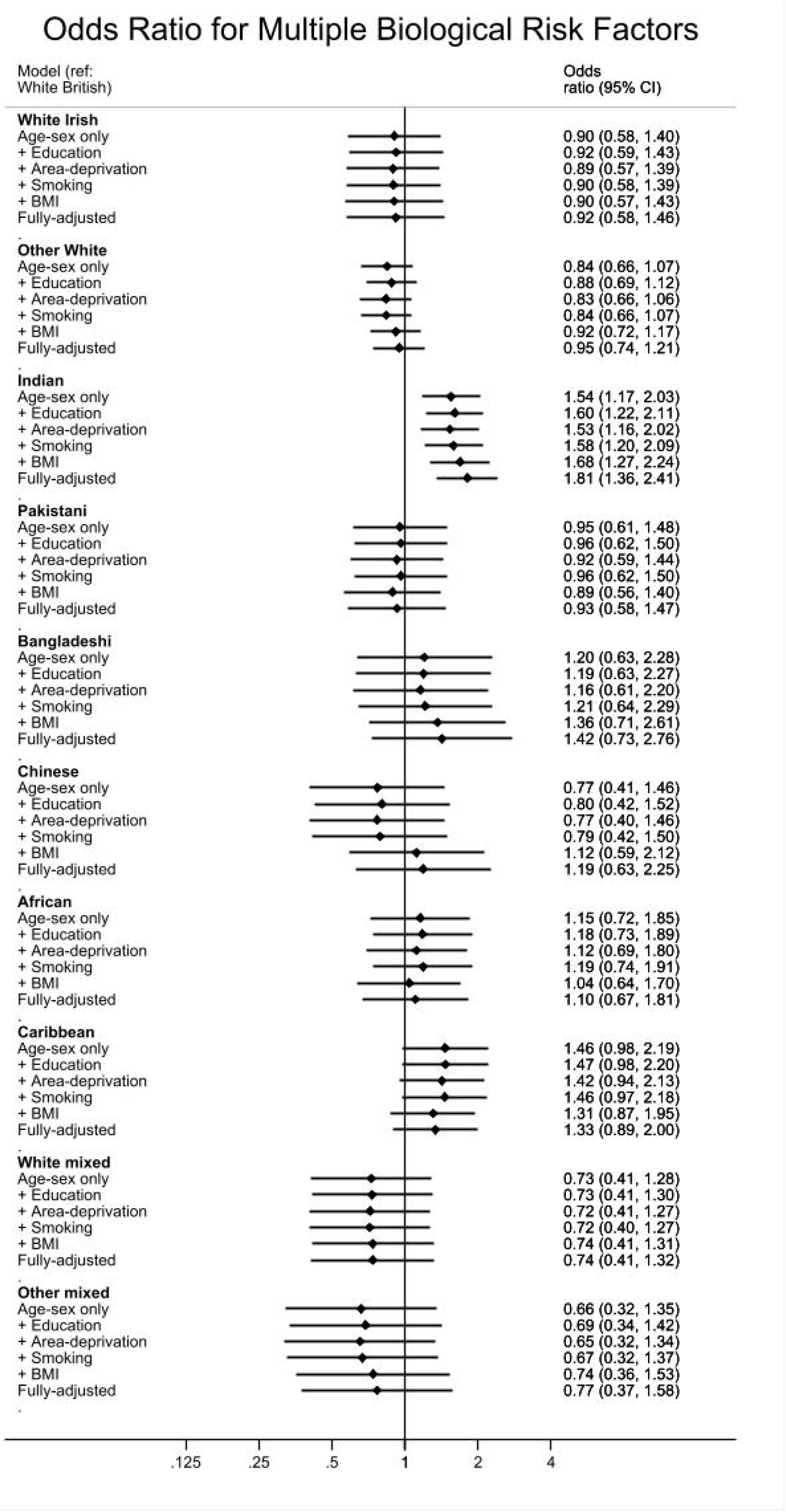
Multiple cardiometabolic risk biomarkers

When based on a larger sample (not restricted to those with BMI measurements), statistically significant results were apparent for similar ethnic groups in the same direction (Table S5). Additionally, Indian adults had lower odds of general multimorbidity (0.78 (0.66-0.91) and Caribbean adults had increased odds of multiple risk biomarkers (1.47 (1.01-2.14)).

## Discussion

General multimorbidity varied with age among ethnic groups, with the risk being greater among the British White population aged under 40, and greater among ethnic minorities (Pakistani and Bangladeshi) aged 40 and over. Age-related differences in patterns of multimorbidity by ethnicity, was also found in a recent study,(22). This correlates well with the increased risk of cardiovascular multimorbidity among Pakistani and Bangladeshi adults aged 40 and over; however, differences between other ethnic groups and greater risk of cardiovascular multimorbidity but not general multimorbidity suggest the general white population has higher prevalence of non-cardiovascular longstanding conditions and/or there are ethnic differences in reporting or in being diagnosed.

Chinese and Asian Indian adults were less likely to report having multimorbidity in the US, however, they were more likely to have a combination of high cholesterol and hypertension, risk factors for CVD consistent with our study,(9). Cultural barriers or differences in considering what constitutes a long-term condition could be a possible explanation for the discrepancy, which requires further investigation. In this study, among those who had cardiovascular multimorbidity, around one in four did not report having general multimorbidity, indicating a discrepancy between definitions, supporting this theory. Healthier ethnic minorities compared with the British White majority, in terms of multimorbidity, could also be due to a ‘healthy migrant effect’,(23). In our study, African, Chinese, Other White and Indian adults (<40 years) had a lower risk of general multimorbidity. Likewise similar groups were found to be less likely than the majority white population to report poorer self-rated health in a different study using the HSE,(15), further highlighting the need to separate these distinct ethnic groups.

Despite the reduced risk of general multimorbidity for some ethnic groups, Indian and African adults and other ethnic groups (Pakistani, Bangladeshi, Caribbean and White Mixed)), were at increased risk of cardiovascular multimorbidity, consistent with a study which found the South Asian and Black populations to have the highest rates of cardiovascular multimorbidity in East London,(4). It is established that ethnic minorities have greater cardiometabolic risk factors,(24). In particular, South Asian and Black adults are more likely to have hypertension and Type 2 Diabetes,(23–25). Moreover, for the South Asian population, the risks of Type 2 diabetes and dyslipidaemia are apparent at lower levels of BMI compared with the white population,(24). We too found Indian and Bangladeshi adults to be at greater risk of cardiometabolic multimorbidity despite having lower levels of obesity than the average in this study. The exact mechanisms behind the differences in risk among ethnicities are not fully understood but are likely due to a complex interplay between genetics and environmental factors,(24).

The association between ethnicity and multimorbidity remained after adjusting for socioeconomic factors, obesity and smoking, suggesting ethnicity to be an independent risk factor for general and cardiovascular multimorbidity. Other studies which adjust for socioeconomic factors have found the greater risk of multimorbidity for ethnic minorities remain,(6,8) or even strengthen, when based on a younger sample,(8). In our models, adjusting for area-deprivation and education tended to reduce the effect, whereas adjustment for smoking and obesity tended to strengthen the effect. In general, after full adjustment, the net effect was a slight reduction in risk, although the Indian population was an exception with a slightly stronger effect after full adjustment, suggesting that the risk for the Indian population is greater despite lower levels of smoking and obesity, and higher levels of educational attainment, which requires further investigation.

Multiple cardiometabolic risk biomarkers exceeding recommended targets, particularly among Indian and Caribbean adults, could be one explanation behind the increase in cardiovascular multimorbidity. Exceeding targets could be exacerbated by undiagnosed diseases or poorer management, such as lack of treatment or adherence to treatment. Ethnic minority groups may face barriers to treatment or health-seeking behaviours. Difficulties with healthcare access are well-recognised,(5). Discrimination, which we were unable to account for, was found to be a risk-factor for multimorbidity in the US,(28) and metabolic syndrome in the Netherlands,(29).

## Strengths and limitations

Strengths of this study include the use of a nationally representative sample in England, where studies based on GP records may only capture people in contact with health services,(5) and studies using a selective sample of volunteers may generate conservative prevalence estimates of multimorbidity,(20). Furthermore, we were able to separate out distinct ethnic groups where differences were found especially in terms of general multimorbidity.

There are many limitations to this study. Even though eight years of data was pooled, sample sizes within distinct ethnic minorities were still relatively small. Consequently, except for cardiovascular multimorbidity, combinations of conditions could not be explored in detail and we were unable to stratify analyses by smaller age groups. We could not adjust for important covariates, such as foreign-born status, which could have suggested a healthy migrant effect, as the question was not asked in the survey years assessed. Furthermore, diet, physical activity, and mental health were not accounted for due to a lack of consistency of data being collected in all years. The extent to which culture, including influences on diet, affect multimorbidity risk could also not be assessed

There is no agreed definition of multimorbidity, and the conditions listed by participants did not include conditions that may be counted in the definition elsewhere, such as alcohol problems,(12) which makes comparing prevalence rates across studies challenging. Finally, our study was cross-sectional therefore we cannot determine the temporal order of events between variables such as deprivation and multimorbidity. However, we believe the use of a nationally representative sample and the ability to assess distinct ethnic groups offset this limitation, where research on ethnicity and multimorbidity is lacking.

## Conclusion

Ethnic inequalities in multimorbidity vary between older and younger adults, distinct ethnic groups, and are independent of social–economic status, smoking and obesity. Differences in findings between general multimorbidity and cardiovascular multimorbidity suggests a possible healthy migrant effect, different susceptibility to different diseases, differential use of healthcare for diagnosis and management, or differences in interpretation of chronic conditions. Several ethnic minority groups among the South Asian and Black ethnic groups are at risk of cardiovascular multimorbidity, which may be exacerbated by underdiagnosis and/or poorer management of cardiometabolic risk factors.

## Data Availability

The Health Survey for England is available to UK Academic Institutions from the UK Data Archive subject to their end-user license.
This study utilised a dataset used for the Health Survey for England report on Ethnicity, which the authors were funded to produce.

http://dx.doi.org/10.5255/UKDA-SN-7260-1

http://dx.doi.org/10.5255/UKDA-SN-7480-1

http://dx.doi.org/10.5255/UKDA-SN-7649-1

http://doi.org/10.5255/UKDA-SN-7919-2

http://doi.org/10.5255/UKDA-SN-8280-1

http://doi.org/10.5255/UKDA-SN-8334-3

http://doi.org/10.5255/UKDA-SN-8488-2

http://doi.org/10.5255/UKDA-SN-8649-1

## Acknowledgements

We thank NHS Digital for funding The Health Survey for England, participants, fieldwork staff and colleagues at Natcen, for collecting, processing the data and creating the dataset

## Contributors

LNF designed and conceived the idea of the study, conducted analyses and drafted the manuscript. JM, LM, SS advised on analyses, and contributed to revisions of the manuscript and approved the final version.

## Ethical approval

Ethical approval was obtained from the following Research Ethics Committees (REC): HSE 2011 and 2012: Oxford A REC: 10/H0604/56; HSE 2013 and 2014: Oxford A REC: 12/sc/0317; HSE 2015: West London NRES Committee: 14/LO/0862; HSE 2016: Nottingham REC: 15/EE/0299, HSE 2017: East of England Research Ethics Committee (Reference no 15/EE/0229),HSE 2018: East Midlands Nottingham 2 Research Ethics Committee (Reference no. 15/EM/0254).. Ethical approval was obtained ahead of data collection, no further ethical approval is needed for secondary analyses.

## Patient consent

Verbal or written consent was obtained from each participant for their involvement ahead of data collection for the various stages of the HSE

## Data availability statement

The Health Survey for England is available to UK Academic Institutions from the UK Data Archive subject to their end-user license. This study utilised a dataset used for the Health Survey for England report on Ethnicity, which the authors were funded to produce.

## Transparency statement

The lead author affirms that this manuscript is an honest, accurate, and transparent account of the study being reported; no important aspects of the study have been omitted; and that any discrepancies from the study as planned have been explained.

